# Differential Vessel features according to the Pathophysiologic Type of Intracranial Atherosclerosis

**DOI:** 10.1101/2023.09.24.23296055

**Authors:** Hyung Jun Kim, Hwan-Ho Cho, Hyunjin Park, Jong-Un Choi, Ha-Na Song, Ji-Eun Lee, In-Young Baek, Jong-Won Chung, Oh Young Bang, Gyeong-Moon Kim, Woo-Keun Seo

## Abstract

**BACKGROUND:** Previous studies have assessed the relationship between vessel tortuosity and atherosclerosis in long vessel segments. We evaluated the relationship between tortuosity and parent artery atherosclerotic disease (PAD) in short vessel segments, and the differences between the PAD and branch atheromatous disease (BAD).

**METHODS:** Computerized analysis of the images provided quantitative vessel features at every 0.2841 mm interval point of the vessel’s centerline. The vessel features that reflect vessel tortuosity include curvature and torsion. To analyze the local features of the vessels, the middle cerebral artery (MCA) was divided into three segments.

**RESULTS:** A total of 59 and 77 patients with PAD and BAD of the MCA were included. Stenotic segments were isolated in 33 patients with PAD and were mostly located in the distal segment; stenotic segments were also isolated in 24 patients with BAD and were mostly located in the middle segment (*P*=0.074). The curvature of the stenotic segment of PAD was significantly increased compared to the non-stenotic segments of the ipsilateral (0.29 ± 0.08 for the stenotic segment vs. 0.26 ± 0.07 for the non-stenotic segment, *P*=0.048), and increased but not significant compared to the stenotic segment of BAD (0.29 ± 0.08 for PAD vs. 0.26 ± 0.05 for BAD, *P*=0.083). In the multivariable regression analysis, curvature (odds ratio, 2.136; 95% confidence interval, 1.251–3.645; *P*=0.005) was associated with PAD.

**CONCLUSIONS:** Our findings suggested that increased tortuosity is associated with the development of PAD and that PAD and BAD have different vessel features and plaque locations and thereby, different pathophysiological mechanisms.

## INTRODUCTION

Intracranial atherosclerosis (ICAS) is one of the most common types of stroke in Asia and South Korea.^1^ The clinical risk factors associated with ICAS include age, race, male sex, hypertension, diabetes mellitus (DM), metabolic syndrome, and dyslipidemia,^2^ and vascular factors, including variations in the circle of Willis and shape of the middle cerebral artery (MCA), have been reported to be associated with the development of atherosclerosis and plaque formation at specific locations.^3,4^

ICAS is classified into parent artery atherosclerotic disease (PAD) and branch atheromatous disease (BAD) based on the characteristics of the ischemic lesion.^5–7^ PAD can be classified into artery-to-artery embolism, thrombotic occlusion due to plaque rupture, and hemodynamic impairment due to severe stenosis.^5^ BAD, which is a subcortical ischemic stroke caused by a parent artery atheroma occluding the orifice of a perforating artery, occurs primarily in the lenticulostriate and anterior pontine arteries.^8^ A study using high-resolution magnetic resonance imaging (HR-MRI) reported different morphological features of plaques in PAD and BAD,^7^ and another study observed an association between different plaque locations in the MCA.^9^ These different plaque features and locations suggest that PAD and BAD have different pathophysiological mechanisms; however, few studies exist on the clinical and vascular factors that influence these pathophysiological mechanisms (Figure 1).^6^

**Figure 1.**
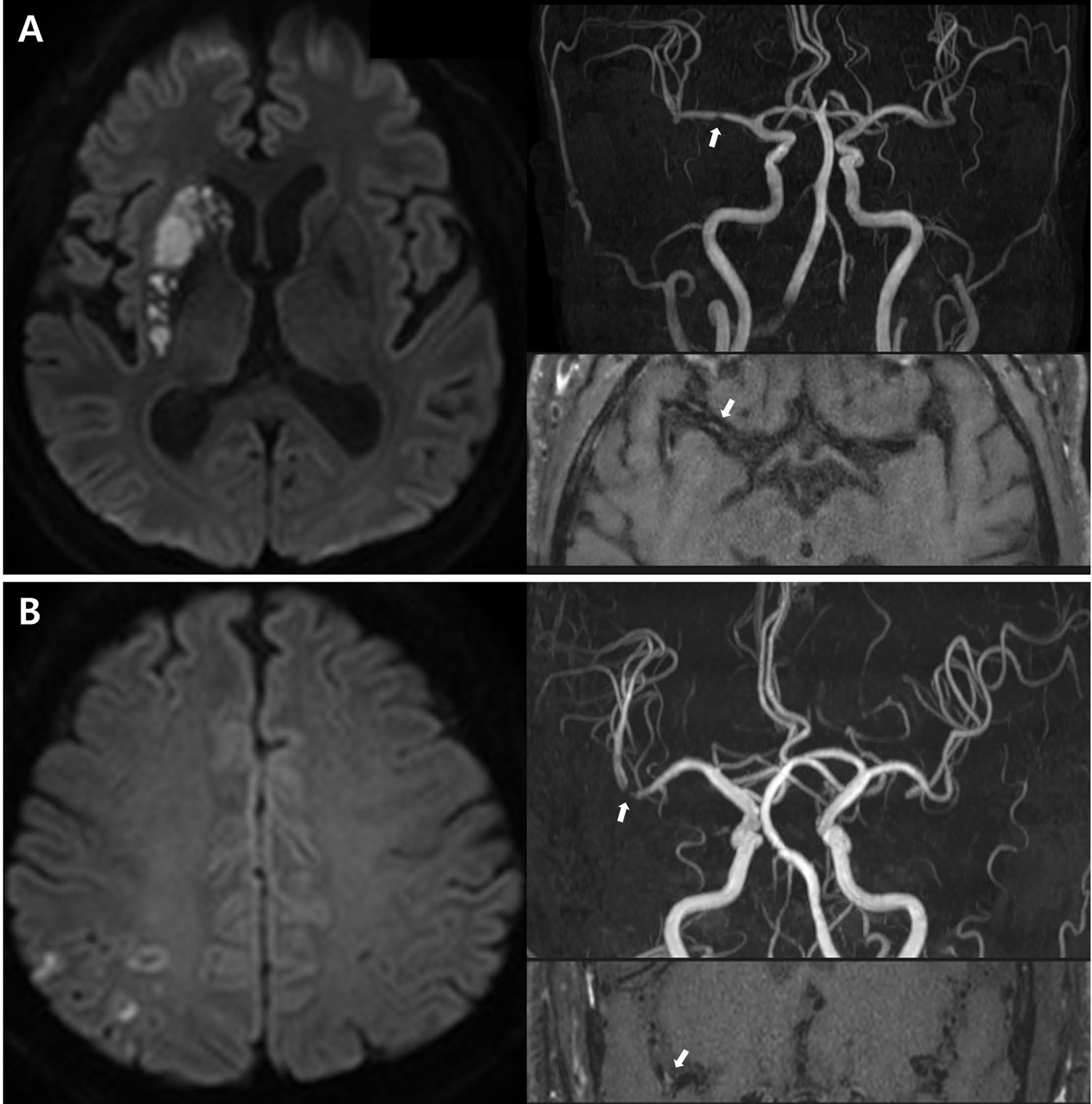
Classification of intracranial atherosclerosis by pathophysiological subtype. (A) Branch atheromatous disease. Diffusion-weighted imaging reveals deep subcortical infarcts. High-resolution magnetic resonance imaging reveals diffuse plaque enhancement with mild stenosis near the orifices of the penetrating arteries (white arrows). (B) Parental atherosclerotic artery disease. Diffusion-weighted image shows small scattered cortical embolic infarcts. Three-dimensional time-of-flight magnetic resonance angiography shows severe stenosis, and high-resolution magnetic resonance imaging shows a plaque in this stenotic area (white arrows).

In the present study, we aimed to analyze the parental vessel features of PAD and BAD to investigate whether different vascular factors are associated with the development of ICAS in different pathophysiological stroke types.

## METHODS

### Study Design and Participants

We retrospectively enrolled consecutive patients with ICAS who were admitted to the Samsung Medical Center (SMC), Seoul, Korea, between January 2017 and December 2019. These patients were included in the SMC stroke registry, which recruits patients with acute stroke within 7 days of stroke onset. All the patients underwent 3-dimensional time-of-flight (3D-TOF) magnetic resonance angiography (MRA) and were enrolled if they had an atherosclerotic plaque protruding into the MCA causing any stenosis (stenosis degree > 0%) in the clinically relevant artery. Some patients with BAD included those with no apparent stenosis on 3D TOF-MRA but with plaques observed on HR-MRI. Patients with PAD included those with stenosis (> 50%) on 3D TOF-MRA, and patients with BAD included those with a lesion size > 20 mm on diffusion MRI and > 0% stenosis on 3D TOF-MRA, or plaques observed on HR-MRI if no obvious stenosis was observed.^10^ Therefore, only patients with the least possible (level of confidence) large-artery atherosclerosis according to the Stop Stroke Study Trial of Org 10172 in Acute Stroke Treatment (SSS-TOAST) classification criteria were enrolled.^11^

To measure the small vessel disease burden of patients in this study, the Fazekas scale scores of all the patients with ICAS was analyzed by two participating neurologists (H. J. K. and W.-K.S.).^12^ To measure the atherosclerotic burden, we quantitatively analyzed the vessels contralateral to the lesion in all the patients and measured the carotid intima-media thickness (carotid IMT) in the middle common carotid artery in 25 patients with PAD and 37 patients with BAD who underwent carotid duplex sonography on admission for stroke management.^13^ The control participants for ICAS were healthy individuals who visited the comprehensive health promotion center at the SMC and underwent MRA between January 1, 2013, and December 31, 2013, excluding those who had the following: (1) stroke including ischemic stroke, hemorrhagic stroke, and transient ischemic attack; (2) coronary artery or heart disease; (3) ICAS; (4) intracranial arterial anomalies corresponding to pathological conditions or variants of normal anatomy; and (5) congenital morbidity including cerebral arterial hypoplasia; and (6) miscellaneous abnormal cases diagnosed by angiography. A total of 136 healthy controls were selected after age- and sex-matching 1:1 with the patients with ICAS. Demographic data and vascular risk factors were collected from the medical records of controls and the stroke registry for the stroke cohort. The Institutional Review Board of SMC approved the study design (SMC-2021-04-072). This study was conducted in accordance with the principles of the Declaration of Helsinki. Informed consent was waived by the SMC Institutional Review Board for the control group because the study progressed in a retrospective manner, and we provided the clinical data and brain images in an anonymized form. Written informed consent was obtained from all the patients enrolled in the SMC stroke registry.

### Imaging Preparation

All the patients underwent routine brain MRI, including 3D-TOF MRA of the intracranial vessels, using a 3-Tesla system (Philips Achieva magnetic resonance imaging scanner). The preprocessing procedures included anonymization using the DICOM Anonymizer Pro (Neologica, Montenotte, Italy) and region growing using an in-house vessel analyzer program to generate fine-grained cerebral angiographic maps and convert them into the NII format.

The internally developed vessel morphology pipelines were analyzed, and brain vessel features were extracted to examine the cerebrovascular structure. Specifically, (i) skeletonizing the cerebrovascular region and surface, (ii) pruning the branch under a predetermined threshold, (iii) generating a linked list of tree structures based on the refined skeletal structure, and (iv) specifying the leaf nodes from the linked list to determine the endpoints. The centerlines were extracted by tracking the boundary surfaces of the cells connecting the start and endpoints. The centerline was connected by points located at 0.2841 mm. Finally, the pipelines characterized numerous blood vessel features of the compartmentalized groups based on the branch point of the centerline. The quantified vessel characteristics included the cerebral blood vessel cross-sectional area, maximum inscribed sphere radius, minimized and maximized diameters, maximum-minimum radius ratio, curvature, torsion, perimeter, luminal circularity, and hydraulic luminal diameter (Table S1). Among these features, the area, maximum inscribed sphere radius, minimized and maximized diameters, maximum-minimum radius ratio, perimeter, luminal circularity, and hydraulic luminal diameter can be changed according to the degree of stenosis. However, the curvature and torsion could be relevant to the vessel tortuosity^14^; thus, further details were provided.

The curvature values were measured as the tangent vector divided by the radius of the osculating circle at each point on the centerline, and torsion values were measured as the change in the osculating plane created by the tangent and normal vectors of the centerline point. Detailed MRI and image analysis protocols have been described previously (Figure 2).^15^

**Figure 2.**
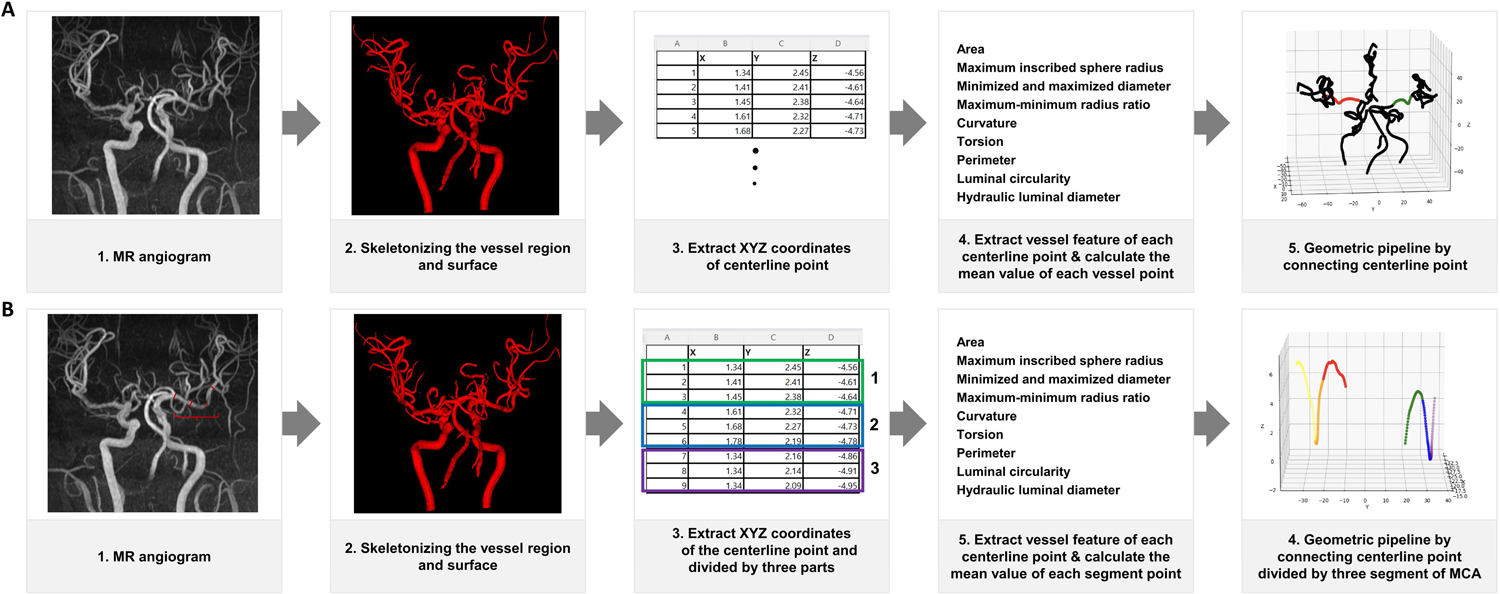
(A) The flow schema describes a comprehensive preprocessing procedure, geometric characterization algorithms, and analysis of the vessel features. (B) Description of the processes for analyzing the middle cerebral artery into three segments.

### Segmentation of the Parent Vessel and Analysis of the Stenotic Segment

To further analyze the local features of the vessels where the plaques were located in the parent artery, the MCA was divided into three segments. In PAD, a segment with stenosis was defined as a stenotic segment, and if multiple segments had stenosis, the segment with the highest stenosis degree was defined as the stenotic segment. In BAD, the stenotic segments were defined as the segment where the plaque was located on HR-MRI. Therefore, patients in whom all the three segments were not visible due to near occlusion or thrombotic occlusion, and those in whom HR-MRI was not performed in BAD were excluded from this analysis. The vessel features of each segment were calculated as the mean values measured at each point (Figure 2).

### Statistical Analysis

Fisher’s exact or chi-square test was used to analyze the categorical variables. Differences in the continuous variables were evaluated using the Student’s t-test and Wilcoxon rank-sum test. The independent factors for PAD development were evaluated using logistic regression analysis. We used two multivariate logistic analysis models: Model 1 included vascular (vessel features of the MCA) and clinical factors influencing the development of PAD, and Model 2 included only vascular factors (curvature and torsion only) influencing the development of PAD (vessel features of divided segments). Vessel features that were attributed to plaque formation rather than influencing plaque development were not used as confounders in the multivariate logistic analysis. A multivariate logistic regression analysis model was used to calculate the odds ratios (OR), 95% confidence intervals (CI), and *p*- values after adjusting for the vascular and clinical factors. All the statistical analyses were performed using the open-source statistical package R version 3.6.3 (R Project for Statistical Computing, Vienna, Austria).

## RESULTS

### General Characteristics

During the study period, 1124 patients were enrolled in the SMC stroke registry; 281 were classified as having large artery atherosclerosis according to the SSS-TOAST classification, and 75 whose parent vessel was not an MCA were excluded. After excluding 54 patients whose vessels could not be analyzed owing to artifacts on the TOF images and 16 patients whose vessels could not be analyzed owing to occlusion from the proximal end of the MCA, 59 patients with PAD and 77 patients with BAD were finally included (Figure 3). A total of 59 patients with PAD (68.25 ± 13.95 years; male, 55.93%), 77 patients with BAD (69.34 ± 11.92 years; male, 59.74%), 59 controls for the patients with PAD (67.49 ± 5.63 years; male, 55.93%), and 77 controls for the patients with BAD (69.34 ± 9.52; male, 59.74%) were included in this study (Table 1). The body mass index (BMI) (PAD vs. BAD: 25 ± 3.44 vs. 23.86 ± 3.13, *P*=0.049) and ischemic stroke history (PAD vs. BAD: 19 [32.2%] vs. 12 [15.58%], *P*=0.037) were significantly higher in PAD compared to BAD (Table 1).

**Figure 3.**
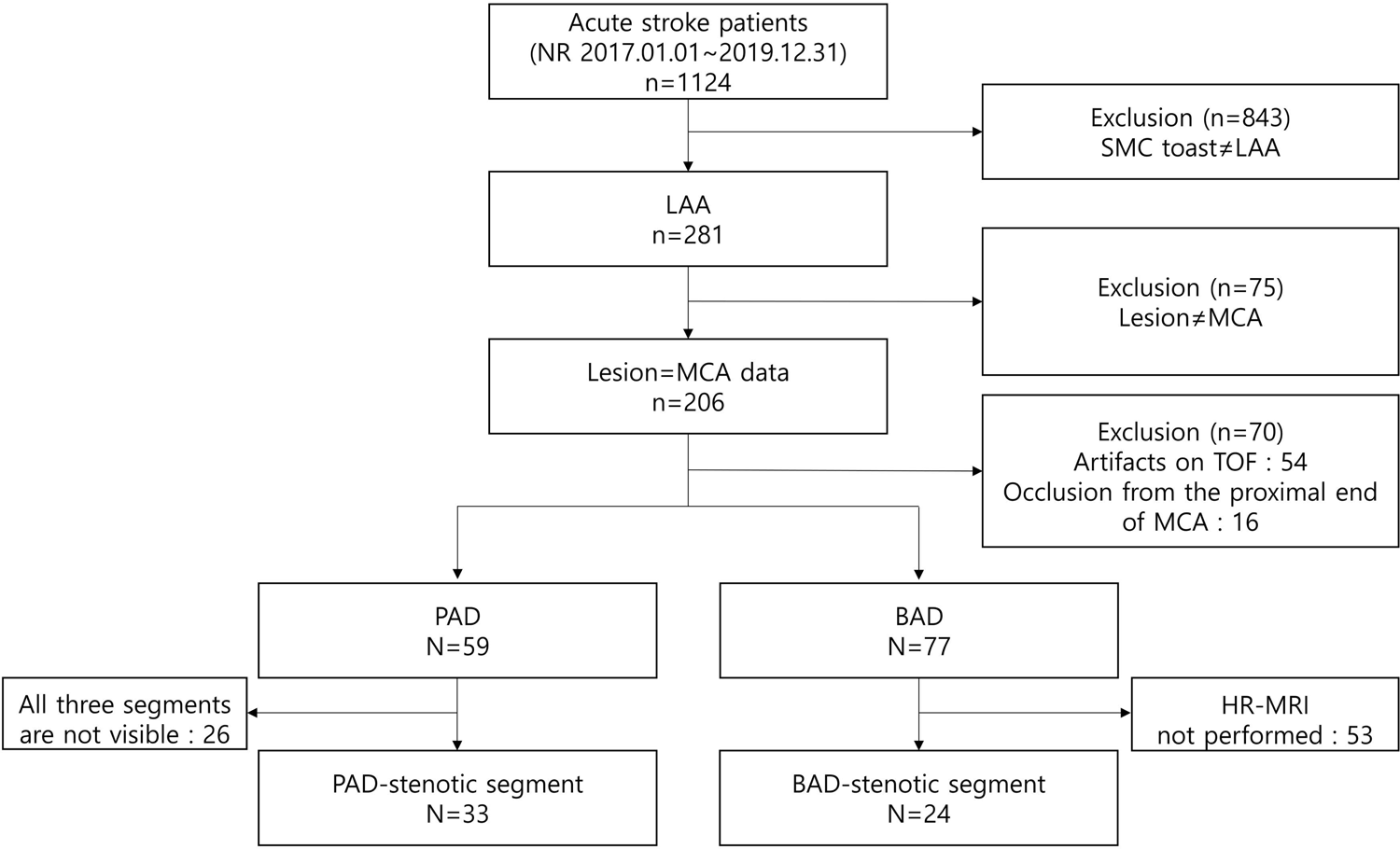
Patient selection.

**Table 1.** Baseline characteristics of the patients with intracranial atherosclerosis.

|  | Parent artery<br>atherosclerotic<br>disease<sup>1</sup> (N=59) | Branch<br>atheromatous<br>disease<sup>2</sup> (N=77) | *P* (1 vs.<br>2) | PAD-control<sup>3</sup><br>(N=59) | BAD-control<sup>4</sup><br>(N=77) | *P*<br>(1 vs. 3) | *P*<br>(2 vs. 4) |
| --- | --- | --- | --- | --- | --- | --- | --- |
| Age, years, mean (SD) | 68.25 ± 13.95 | 69.34 ± 11.92 | 0.634 | 67.49 ± 5.63 | 69.34 ± 9.52 | 0.698 | 0.921 |
| Male, (%) | 33 (55.93%) | 46 (59.74%) | 0.787 | 33 (55.93%) | 46 (59.74%) | >0.99 | >0.99 |
| BMI (kg/m<sup>2</sup>), mean (SD) | 25 ± 3.44 | 23.86 ± 3.13 | 0.049 | 23.5 ± 2.63 | 24.74 ± 2.97 | 0.009 | 0.074 |
| **Vascular risk factors** |  |  |  |  |  |  |  |
| Hypertension, (%) | 49 (83.05%) | 61 (79.22%) | 0.732 | 23 (38.98%) | 25 (32.47%) | <0.001 | <0.001 |
| Diabetes mellitus, (%) | 24 (40.68%) | 19 (24.68%) | 0.071 | 9 (15.25%) | 13 (16.88%) | 0.004 | 0.321 |
| Hypercholesterolemia, (%) | 40 (67.8%) | 59 (76.62%) | 0.341 | 23 (38.98%) | 26 (33.77%) | 0.003 | <0.001 |
| Current smoking, (%) | 13 (22.03%) | 13 (16.88%) | 0.697 | 3 (5.08%) | 8 (10.39%) | 0.016 | 0.348 |
| **Comorbidities** |  |  |  |  |  |  |  |
| Previous ischemic stroke, (%) | 19 (32.2%) | 12 (15.58%) | 0.037 | N/A | N/A | N/A | N/A |
| Coronary disease, (%) | 7 (8.97%) | 8 (7.34%) | 0.219 | N/A | N/A | N/A | N/A |
| **Medication before admission** |  |  |  |  |  |  |  |
| Previous antiplatelet, (%) | 21 (35.59%) | 23 (29.87%) | 0.602 | N/A | N/A | N/A | N/A |
| Previous statin, (%) | 22 (37.29%) | 22 (28.57%) | 0.372 | N/A | N/A | N/A | N/A |
| **Initial NIHSS, (IQR)** | 3 (1, 6) | 3 (2, 4) | 0.780 | N/A | N/A | N/A | N/A |
| **END, (%)** | 0 (0%) | 10 (12.99%) | 0.011 | N/A | N/A | N/A | N/A |
| **Laboratory test** |  |  |  |  |  |  |  |
| LDL (mg/dL) | 118.88 ± 39.66 | 128.3 ± 43.21 | 0.189 | N/A | N/A | N/A | N/A |
| TG (mg/dL) | 173.31 ± 109.78 | 174.53 ± 114.63 | 0.950 | 102.92 ± 44.67 | 122 ± 64.78 | <0.001 | <0.001 |
| Fasting glucose (mg/dL) | 123.26 ± 38.98 | 117.42 ± 45.3 | 0.428 | 102.78 ± 20.44 | 104.18 ± 21.63 | <0.001 | 0.025 |
| Homocysteine (mg/dL) | 13.86 ± 6.64 | 12.76 ± 5.28 | 0.320 | N/A | N/A | N/A | N/A |
| **Carotid Intima-Media thickness** |  |  |  |  |  |  |  |
| Right (mm) | 1.58 ± 0.97 | 1.49 ± 0.87 | 0.718 |  |  |  |  |
| Left (mm) | 1.46 ± 0.88 | 1.33 ± 0.64 | 0.523 |  |  |  |  |
| **Fazekas scale** |  |  |  |  |  |  |  |
| Periventricular lesions (IQR) | 2 (1, 2) | 2 (1, 2) | 0.925 |  |  |  |  |
| White matter lesions (IQR) | 1 (1, 2) | 1 (1, 2) | 0.410 |  |  |  |  |

**PAD, parent artery atherosclerotic disease; BAD, branch atheromatous disease; SD, standard deviation; BMI, body mass index; NIHSS, national institutes of health stroke scale; END, early neurological deterioration; LDL, low-density lipoprotein; TG, triglyceride; IQR, interquartile range**

### Vessel Feature According to the ICAS Subtypes

Minimum-maximum radius ratio (PAD vs. BAD: 0.81 ± 0.04 vs. 0.83 ± 0.04, *P*=0.002) and luminal circularity (PAD vs. BAD: 0.95 ± 0.02 vs. 0.96 ± 0.02, *P*=0.022) were higher in the BAD than in the PAD cases. Compared to the PAD control, PAD was significantly lower in area (PAD vs. PAD-control: 11.28 ± 3.04 vs. 12.43 ± 2.29, *P*=0.022), maximum inscribed sphere radius (PAD vs. PAD-control: 1.62 ± 0.21 vs. 1.79 ± 0.13, *P*<0.001), minimum diameter (PAD vs. PAD-control: 3.32 ± 0.46 vs. 3.62 ± 0.26, *P*<0.001), minimum-maximum diameter ratio (PAD vs. PAD-control: 0.81 ± 0.04 vs. 0.85 ± 0.04, *P*<0.001), perimeter (PAD vs. PAD-control: 12.19 ± 1.85 vs. 12.85 ± 1.42, *P*=0.034), luminal circularity (PAD vs. PAD- control: 0.95 ± 0.02 vs. 0.96 ± 0.02, *P*=0.004), and hydraulic-luminal diameter (PAD vs. PAD-control: 3.52 ± 0.4 vs. 3.78 ± 0.29, *P*<0.001), and significantly higher in curvature (PAD vs. PAD-control: 0.27 ± 0.04 vs. 0.23 ± 0.03, *P*<0.001). However, compared to the BAD control, BAD was significantly lower in area (BAD vs. BAD-control: 11.43 ± 2.13 vs. 12.55 ± 2.23, *P*=0.002), maximum inscribed sphere radius (BAD vs. BAD-control: 1.68 ± 0.16 vs. 1.80 ± 0.15, *P*<0.001), minimum diameter (BAD vs. BAD-control: 3.42 ± 0.35 vs. 3.65 ± 0.33, *P*<0.001), maximum diameter (BAD vs. BAD-control: 4.25 ± 0.47 vs. 4.42 ± 0.49, *P*=0.033), minimum-maximum radius ratio (BAD vs. BAD-control: 0.83 ± 0.04 vs. 0.85 ± 0.04, *P*=0.001), perimeter (BAD vs. BAD-control: 12.30 ± 1.28 vs. 12.89 ± 1.29, *P*=0.005), and hydraulic-luminal diameter (BAD vs. BAD-control: 3.60 ± 0.33 vs. 3.8 ± 0.31, *P*<0.001), and significantly higher in curvature (BAD vs. BAD-control: 0.26 ± 0.03 vs. 0.24 ± 0.04, *P*<0.001) (Table 2).

**Table 2.** Comparison of the vessel features between the parent artery atherosclerotic disease and branch atheromatous disease, and normal controls.

|  | Total (N=136) | Parent artery<br>atherosclerotic<br>disease<sup>1</sup> (N=59) | Branch<br>atheromatous<br>disease<sup>2</sup> (N=77) | PAD-control<sup>3</sup><br>(N=59) | BAD-control<sup>4</sup><br>(N=77) | *P* (1 vs<br>2) | *P* (1 vs<br>3) | *P* (2 vs<br>4) |
| --- | --- | --- | --- | --- | --- | --- | --- | --- |
| Area | 11.36 ± 2.55 | 11.28 ± 3.04 | 11.43 ± 2.13 | 12.43 ± 2.29 | 12.55 ± 2.23 | 0.751 | 0.022 | 0.002 |
| Max.InsRad | 1.66 ± 0.19 | 1.62 ± 0.21 | 1.68 ± 0.16 | 1.79 ± 0.13 | 1.8 ± 0.15 | 0.076 | <0.001 | <0.001 |
| Min. | 3.38 ± 0.4 | 3.32 ± 0.46 | 3.42 ± 0.35 | 3.62 ± 0.26 | 3.65 ± 0.33 | 0.157 | <0.001 | <0.001 |
| Max. | 4.25 ± 0.56 | 4.25 ± 0.67 | 4.25 ± 0.47 | 4.42 ± 0.52 | 4.42 ± 0.49 | 0.974 | 0.123 | 0.033 |
| MaxMinRat | 0.82 ± 0.04 | 0.81 ± 0.04 | 0.83 ± 0.04 | 0.85 ± 0.04 | 0.85 ± 0.04 | 0.002 | <0.001 | 0.001 |
| Curvature | 0.26 ± 0.04 | 0.27 ± 0.04 | 0.26 ± 0.03 | 0.23 ± 0.03 | 0.24 ± 0.04 | 0.362 | <0.001 | <0.001 |
| Torsion | 2.08 ± 0.53 | 2.03 ± 0.27 | 2.12 ± 0.67 | 2.09 ± 0.34 | 2.12 ± 0.43 | 0.283 | 0.329 | 0.968 |
| Perimeter | 12.25 ± 1.55 | 12.19 ± 1.85 | 12.3 ± 1.28 | 12.85 ± 1.42 | 12.89 ± 1.29 | 0.706 | 0.034 | 0.005 |
| Luminal circularity | 0.96 ± 0.02 | 0.95 ± 0.02 | 0.96 ± 0.02 | 0.96 ± 0.02 | 0.96 ± 0.02 | 0.022 | 0.004 | 0.336 |
| Hydr. | 3.57 ± 0.39 | 3.52 ± 0.4 | 3.6 ± 0.33 | 3.78 ± 0.29 | 3.8 ± 0.31 | 0.228 | <0.001 | <0.001 |

**PAD**, parent artery atherosclerotic disease; **BAD**, branch atheromatous disease; **Max.InsRad**, Maximum inscribed sphere radius; **Min.**, Minimized diameter; **Max.**, Maximized diameter; **Max-Min ratio**, Maximum-minimum radius ratio; **Hydr.**, Hydraulic luminal diameter

### Vessel Feature of the Stenotic Segment vs. Non-Stenotic Segment

The stenotic segment was isolated from 33 patients with PAD and was most commonly located in the distal segment (proximal segment,5 [15.15%]; middle segment,7 [21.21%]; and distal segment, 21 [63.64%]). The maximum inscribed sphere radius was significantly lower (1.44 ± 0.31 in the stenotic segment vs. 1.6 ± 0.34 in the non-stenotic segment; *P*=0.020), and the curvature of the stenotic segment was significantly higher (0.29 ± 0.08 for the stenotic segment vs. 0.26 ± 0.07 for the non-stenotic segment, *P*=0.048) in the stenotic segments compared to the non-stenotic segments. Compared to the contralateral part of the stenotic segment (PAD-contralateral), the stenotic segment was significantly lower in area (9.19 ± 4 for the stenotic segment vs. 12.72 ± 2.83 for the contralateral segment, *P*<0.001), maximum inscribed sphere radius (1.44 ± 0.31 for the stenotic segment vs. 1.74 ± 0.19 for the contralateral segment, *P*<0.001), minimum diameter (2.98 ± 0.67 for the stenotic segment vs. 3.55 ± 0.41 for the contralateral segment, *P*<0.001), maximum diameter (3.77 ± 0.96 for the stenotic segment vs. 4.57 ± 0.66 for the contralateral segment, *P*<0.001), perimeter (10.81 ± 2.47 for the stenotic segment vs. 13.04 ± 1.72 for the contralateral segment, *P*<0.001), and hydraulic-luminal diameter (3.19 ± 0.73 for the stenotic segment vs. 3.74 ± 0.38 for the contralateral segment, *P*<0.001), and significantly higher in curvature (0.29 ± 0.08 for the stenotic segment vs. 0.25 ± 0.04 for the contralateral segment, *P*=0.026).

The stenotic segment was isolated from 24 patients with BAD and was most commonly located in the middle segment (proximal segment, 2 [8.33%]; middle segment, 12 [50.00%]; and distal segment, 10 [41.67%]). Patients with BAD had no vessel features that were significantly different between the stenotic and non-stenotic segments. However, the comparison between the stenotic and contralateral segment showed differences in area (10.03 ± 2.69 for the stenotic segment vs. 12.05 ± 2.58 for the contralateral segment, *P*=0.012), maximum inscribed sphere radius (1.59 ± 0.22 for the stenotic segment vs. 1.74 ± 0.18 for the contralateral segment, *P*=0.011), minimum diameter (3.25 ± 0.46 for the stenotic segment vs. 3.54 ± 0.41 for the contralateral segment, *P*=0.028), maximum diameter (3.93 ± 0.61 for the stenotic segment vs. 4.38 ± 0.52 for the contralateral segment, *P*=0.010), perimeter (11.4 ± 1.63 for the stenotic segment vs. 12.7 ± 1.46 for the contralateral segment, *P*=0.007) and hydraulic-luminal diameter (3.41 ± 0.46 for the stenotic segment vs. 3.71 ± 0.39 for the contralateral segment, *P*=0.020).

In the analysis comparing the stenotic segment of PAD with the stenotic segment of BAD, only the maximum inscribed sphere radius (1.44 ± 0.31 for the stenotic segment of PAD vs. 1.59 ± 0.22 for the stenotic segment of BAD, *P*=0.040) was significantly lower in PAD compared to BAD; although not significant, a trend of difference in plaque location was observed (*P*=0.074). Curvature (0.29 ± 0.08 for the stenotic segment of PAD vs. 0.26 ± 0.05 for the stenotic segment of BAD, *P*=0.083) was also observed to be higher in PAD (Table 4).

### Factors Associated With PAD

After adjusting for the BMI and DM in the multivariate logistic regression analysis (Model 1), BMI (OR, 1.132; 95% CI, 1.011–1.268; *P*=0.032) was associated with PAD in ICAS (Table 3). Moreover, in the analysis of vessel features associated with PAD development (Model 2), curvature (OR, 2.136; 95% CI, 1.251–3.645; *P*=0.005) was associated with PAD in the multivariable logistic regression analysis adjusted for curvature and torsion (Table 5).

**Table 3.** Vascular and clinical factors associated with the parent artery atherosclerotic disease in 136 patients with intracranial atherosclerosis stroke.

|  | Univariate | p | Multivariate | p |
| --- | --- | --- | --- | --- |
| **Curvature**<br>**(mean value of M1)** | 0.766 (0.546–1.076) | 0.124 |  |  |
| **Torsion**<br>**(mean value of M1)** | 0.647 (0.252–1.661) | 0.365 |  |  |
| **Age, years** | 0.766 (0.546–1.076) | 0.124 |  |  |
| **Male** | 0.855 (0.430–1.670) | 0.656 |  |  |
| **BMI (kg/m<sup>2</sup>)** | 1.114 (1.001–1.241) | 0.049 | 1.132 (1.011–1.268) | 0.032 |
| **Hypertension** | 1.285 (0.536–3.083) | 0.574 |  |  |
| **Diabetes mellitus** | 2.093 (1.005–4.360) | 0.048 |  |  |
| **Hypercholesterolemia** | 0.642 (0.3001–1.373) | 0.253 |  |  |
| **Current smoking** | 1.391 (0.591–3.278) | 0.450 |  |  |
| **LDL (mg/dL)** | 0.995 (0.986–1.003) | 0.194 |  |  |
| **TG (mg/dL)** | 1.000 (0.997–1.003) | 0.949 |  |  |
| **Fasting glucose (mg/dL)** | 1.003 (0.995–1.011) | 0.436 |  |  |
| **Homocysteine (mg/dL)** | 1.032 (0.971–1.097) | 0.305 |  |  |
| **BMI, body mass index; LDL, low-density lipoprotein; TG, triglyceride** |  |  |  |  |

**Table 4.**
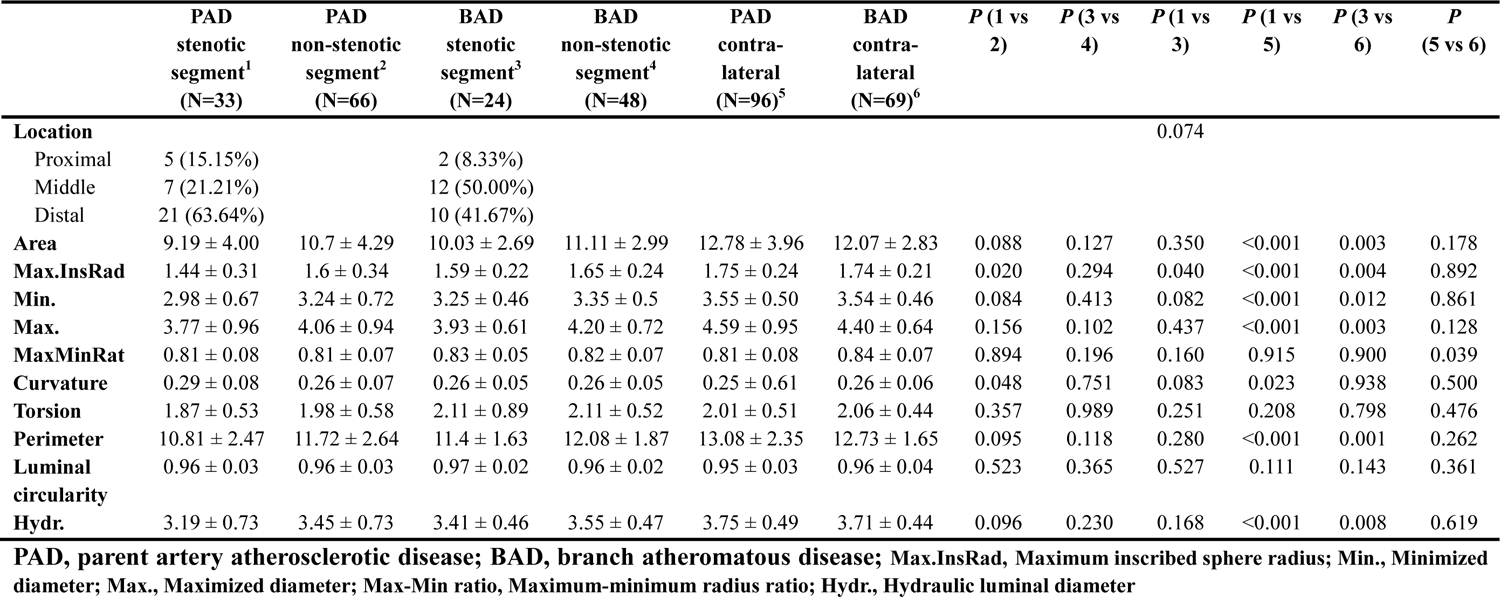
Comparison of the vessel feature between the stenotic and non-stenotic segment of parent artery atherosclerotic disease, stenotic and non-stenotic segment of branch atheromatous disease, and contralateral to the lesion.

**Table 5.** Vascular factors associated with the parent artery atherosclerotic disease in 336 segments of middle cerebral artery from 57 patients with intracranial atherosclerosis.

|  | Univariate | *P* | Multivariate | *P* |
| --- | --- | --- | --- | --- |
| **Area** | 0.817 (0.731–0.914) | <0.001 |  |  |
| **Max.InsRad** | 0.047 (0.012–0.176) | <0.001 |  |  |
| **Min.** | 0.285 (0.153–0.531) | <0.001 |  |  |
| **Max.** | 0.441 (0.274–0.710) | <0.001 |  |  |
| **MaxMinRat** | 0.164 (0.002–17.905) | 0.450 |  |  |
| **Curvature** | 2.112 (1.237–3.608) | 0.006 | 2.136 (1.251–3.645) | 0.005 |
| **Torsion** | 0.518 (0.239–1.121) | 0.095 | 0.491 (0.220–1.098) | 0.083 |
| **Perimeter** | 0.721 (0.608–0.857) | <0.001 |  |  |
| **Luminal circularity** | 24.123 (0.001–357.608) | 0.417 |  |  |
| **Hydr.** | 0.307 (0.168–0.563) | <0.001 |  |  |
| **Location (ref. proximal)** |  |  |  |  |
| **Middle** | 1.322 (0.407–4.299) | 0.642 |  |  |
| **Distal** | 4.607 (1.670–12.710) | 0.003 |  |  |

Max.InsRad, Maximum inscribed sphere radius; Min., Minimized diameter; Max., Maximized diameter; Max-Min ratio, Maximum-minimum radius ratio; Hydr., Hydraulic luminal diameter

## DISCUSSION

This study explored the role of the morphological features of the cerebral artery in the mechanism of stroke and demonstrated that tortuosity, measured by the curvature of the diseased arterial segment, was associated with PAD. In patients with PAD, the target arterial segment tends to be located in the distal part of the MCA. ICAS has two pathophysiological mechanisms: PAD and BAD. The terms PAD and BAD were originally created based on pathological findings; however, few pathological studies have been conducted to date.^16^ With the availability of new generations of computed tomography scanners and MRI technology, the concept of BAD was added to ICAS,^17^ and various studies on PAD and BAD based on neuroimaging have since been conducted.^10^ Recently, the availability of HR-MRI has made it possible to observe the morphological and pathological changes of plaques in the walls of intracranial vessels.^7,18^ These studies have revealed differences in the plaque morphology and location between PAD and BAD in the MCA, suggesting different pathophysiological mechanisms of plaque development.^7,9^ However, research on the vascular factors that contribute to the development of different morphological plaques at different locations in ICAS with distinct pathophysiological mechanisms is lacking. This could be attributed to the fact that the vessel features of cerebral arteries have not been easy to quantify until now, and measuring short segmental vessel tortuosity with methods used in previous studies is challenging.^19^ A point every 0.2841 mm on the centerline of the vessel was established and the vessel features for each point was quantified in our study; therefore, measuring the vessel features of a short segment was possible. The curvature value can be measured for each point, and the mean value of the curvature at a particular segment can reflect vessel tortuosity.^20^ In our study, we quantified the vessel features in specific segments where plaques were located and analyzed the vessel features associated with the development of PAD.

Tortuosity refers to the abnormal twisting, curving, or bending of blood vessels, particularly arteries.^21^ Atherosclerosis is a chronic inflammatory disease characterized by the development of plaques within arterial walls.^22^ Although no direct causal relationship exists between tortuosity and atherosclerosis,^21^ several associations and interactions between them have been reported.^23,24^ Tortuous vessels can cause alterations in the blood flow patterns, including changes in the velocity and turbulence.^25^ This change in blood flow resulting from tortuosity can cause low oscillatory shear stress in certain regions of the vessel, which is associated with endothelial dysfunction and increased susceptibility to atherosclerotic plaque development.^23,25,26^ However, certain coronary artery studies have reported a negative correlation between tortuosity and atherosclerosis,^27–30^ whereas other coronary and cerebral artery studies have reported a positive correlation.^31,32^ Because intra-cranial vessels have a different vessel wall composition than extra-cranial vessels, and the paraclinoid internal carotid artery has greater tortuosity, which makes it more prone to calcification than other intracranial vessels,^2,33^ it is reasonable to assume a possible association between tortuosity and atherosclerosis in intra-cranial vessels.^34^ Plaque formation with low or oscillatory low wall shear stress located along the internal curvature is the most susceptible to plaque formation; moreover, severe tortuosity is likely to be more susceptible to plaque formation.^26,35^ In our study, the curvature of a particular MCA segment, which represents the degree of tortuosity, was associated with PAD, suggesting that tortuosity is linked to shear stress and may contribute to PAD development.

In contrast, BAD has a diffuse atherosclerotic plaque that is distinct from PAD, which occludes the orifices of the small branches.^6^ BAD is often mistaken as a milder form of PAD owing to the lesser degree of stenosis or as a small artery occlusion due to the pattern of ischemic lesions on diffusion images.^10^ However, previous studies have demonstrated that the non-relevant intra- and extra-cranial atherosclerosis burden in patients with BAD did not differ from that in patients with PAD^6^; moreover, another study demonstrated that BAD has fewer small vessel disease features, such as leukoaraiosis and microbleeds, than small artery occlusion.^36^ In addition, unlike small artery occlusion, BAD is more prone to early neurological deterioration due to the presence of plaque in the parent artery surrounding the branch orifice, which can gradually progress from near occlusion to total occlusion, or an unstable plaque near the orifice can cause distal embolization.^37^ In the present study, no differences were observed in the vessel features of the contralateral vessels and the carotid IMT. In addition, we measured the Fazekas scale scores in all the patients with ICAS and found no difference in the small-vessel disease burden between the two ICAS subtypes. We found no significant differences in the atherosclerotic and small vessel disease burden between different ICAS subtypes, supporting previous studies and our hypothesis that BAD is not a milder form of PAD but is caused by a different pathophysiological mechanism.^6^ Furthermore, in terms of clinical characteristics, including vascular risk factors, BAD is more similar to PAD than to small artery disease,^8,9^ and in our study, only BMI was significantly different among the clinical factors. Therefore, local vascular rather than systemic clinical factors may contribute to the different pathophysiological mechanisms of PAD and BAD. Very little research exists on the vascular factors that cause the development of diffuse plaques around the orifice of the branch compared to PAD. Wall shear stress is also lower around branches; thus, it can be assumed that these hemodynamic stresses act around the orifice to cause endothelial dysfunction, leading to plaque development.^26^ Further hemodynamic studies are warranted to determine the factors that lead to differently shaped plaques at different locations.

This study had several limitations. First, this was a single-center study of Koreans, and the results should be interpreted with caution to the general population and other ethnicities. Further multicenter studies involving other ethnicities are needed. Second, the analysis that divided the MCA into three segments included only patients who underwent HR-MRI; therefore, the possibility of selection bias exists. Future studies should be conducted in consecutive patients with BAD undergoing HR-MRI. Third, we analyzed the carotid IMT to measure the atherosclerotic burden; however, because we enrolled patients retrospectively, carotid duplex sonography was not performed in all the patients. Therefore, the possibility of selection bias exists, which may have affected our results. However, to compensate for this limitation, we also analyzed the vessel features contralateral to the lesion and found no differences between the ICAS subtypes. Fourth, although this is a study in a longitudinal study and demonstrated an association between vessel tortuosity and PAD, causality was not established. Therefore, caution should be exercised when interpreting these results.

## CONCLUSION

Our results demonstrated that an increase in the vessel tortuosity was associated with the development of PAD in patients with ICAS. Our study also suggests that PAD and BAD in the MCA have different vessel features and plaque locations and thus have different pathophysiological mechanisms based on the ICAS subtype.

## Data Availability

The data presented in this study are available on request from the corresponding authors. The data are not publicly available due to privacy.

## Author contributions

H.J. Kim: Study concept and design, analysis and interpretation of data, drafting/revision of the manuscript for content, and statistical analysis. H.-N. Song, J.-E. Lee and I.-Y. Baek: Data acquisition, analysis and interpretation J.-W. Chung, O.Y., Bang, G.-M. Kim: Study conceptualization and design, data analysis, and interpretation W.-K. Seo: Study conceptualization and design; data acquisition, analysis, and interpretation;, and drafting/revision of the manuscript for content.

## Acknowledgments

None

